# Air Quality and COVID-19 Prevalence/Fatality

**DOI:** 10.1101/2020.06.14.20130740

**Authors:** Hisato Takagi, Toshiki Kuno, Yujiro Yokoyama, Hiroki Ueyama, Yosuke Hari, Tomo Ando

## Abstract

To investigate the association of real-time/observed ozone/PM2.5 levels with COVID-19 prevalence/fatality, meta-regression of data from the Northeast megalopolis was conducted. Daily Air Quality Index (AQI) values based on available ozone/PM2.5 data in these counties/cities (3/15/2020–5/31/2020) were extracted from US Environmental Protection Agency and World Air Quality Project. In each county/city, total confirmed COVID-19 cases/deaths (5/31/2020) were available from Johns Hopkins Coronavirus Resource Center, and total population was extracted from US Census Bureau. Random-effects meta-regression was performed using OpenMetaAnalyst. A meta-regression graph depicted COVID-19 prevalence and fatality (plotted as logarithm-transformed prevalence/fatality on the y-axis) as a function of mean ozone/PM2.5 AQI (plotted on the x-axis). Coefficients were not statistically significant for ozone (P = 0.212/0.814 for prevalence/fatality) and PM2.5 (P = 0.986/0.499). Although multivariable analysis had been planned, it was not performed because of non-significant covariates of interest in the univariable model. In conclusion, ozone/PM2.5 may be unassociated with COVID-19 prevalence/fatality.

## Introduction

Air quality defined by ozone/particulate matter 2.5 (PM2.5)/PM10/CO/SO2/NO2/etc. has been known to affect pulmonary/cardiac diseases. In the recent coronavirus disease 2019 (COVID-19) pandemic, a few studies^1-4^ suggest that air quality also may militate against COVID-19 prevalence/case fatality. To investigate the association of real-time/observed (neither historical nor estimated) ozone/PM2.5 levels with COVID-19 prevalence/fatality, meta-regression (considering a county/city as a study in meta-analysis and weighted by inverse variance of prevalence/fatality) of data from the Northeast megalopolis (including >50-million folks, >15% of US total population) was conducted.

## Methods

Five Combined Statistical Areas in the Northeast megalopolis comprise 111 counties/cities. Daily Air Quality Index (AQI) values based on available ozone/PM2.5 data in these counties/cities (3/15/2020, beginning of rapid increase of new COVID-19 cases, to 5/31/2020) were extracted from US Environmental Protection Agency (https://www.epa.gov/outdoor-air-quality-data/air-data-daily-air-quality-tracker) and World Air Quality Project (https://aqicn.org/city/usa/newyork/). In each county/city, total confirmed COVID-19 cases/deaths (5/31/2020) were available from Johns Hopkins Coronavirus Resource Center (https://github.com/CSSEGISandData/COVID-19/tree/master/csse_covid_19_data/csse_covid_19_daily_reports), and total population was extracted from US Census Bureau (https://www.census.gov/acs/www/data/data-tables-and-tools/data-profiles/). Random-effects meta-regression was performed using OpenMetaAnalyst (http://www.cebm.brown.edu/openmeta/index.html). A meta-regression graph depicted COVID-19 prevalence (defined as total cases divided by total population) and fatality (defined as total deaths divided by total cases) (plotted as logarithm-transformed prevalence/fatality on the y-axis) as a function of mean ozone/PM2.5 AQI during 3/15/2020–5/31/2020 (plotted on the x-axis).

## Results

Coefficients were not statistically significant for ozone (P = 0.212/0.814 for prevalence/fatality; Fig. 1A/1B) and PM2.5 (P = 0.986/0.499; Fig. 1C/1D), which would indicate that COVID-19 prevalence/fatality was unaffected by ozone/PM2.5. Although multivariable analysis had been planned, it was not performed because of non-significant covariates of interest in the univariable model.

**Figure 1.**
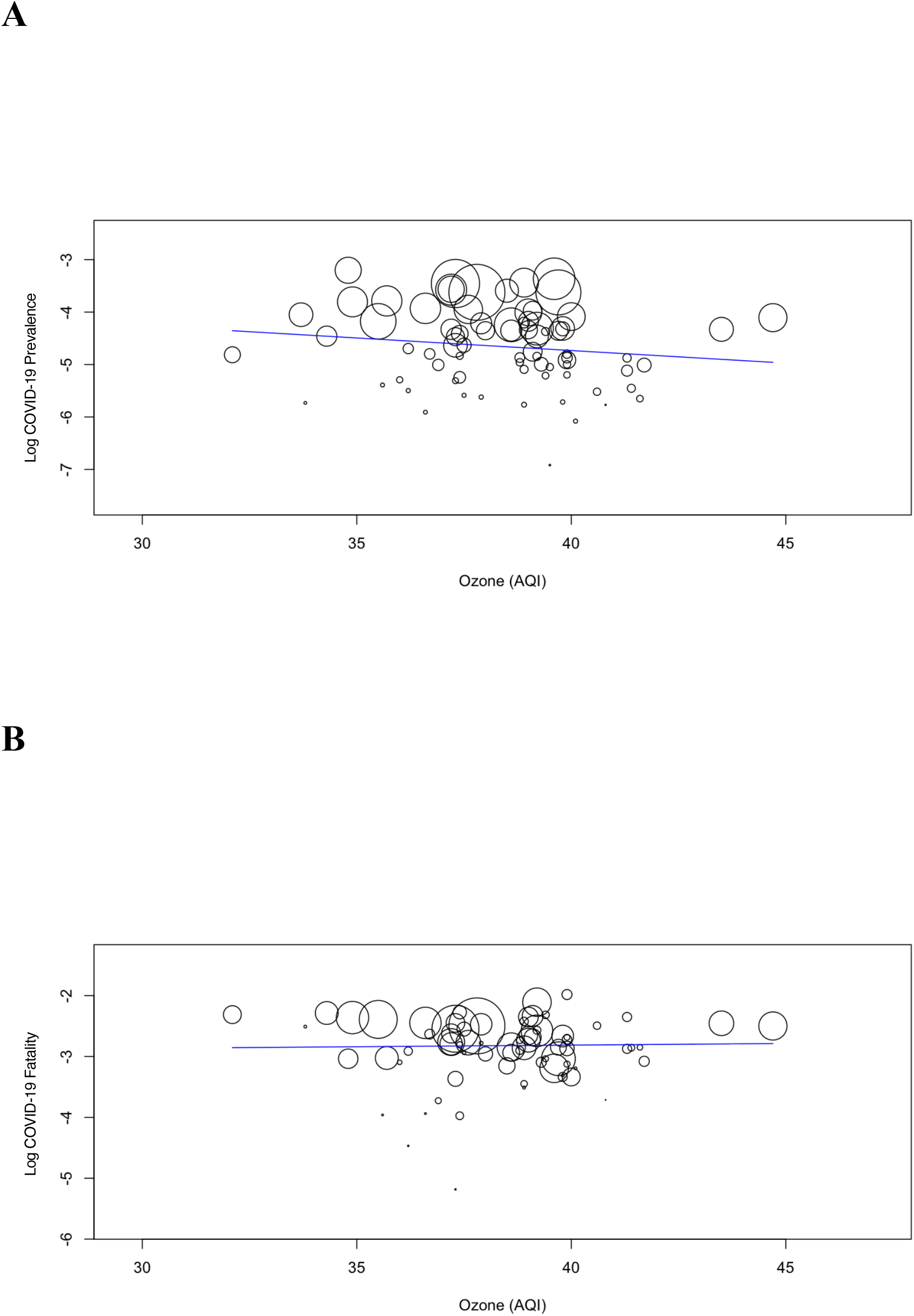
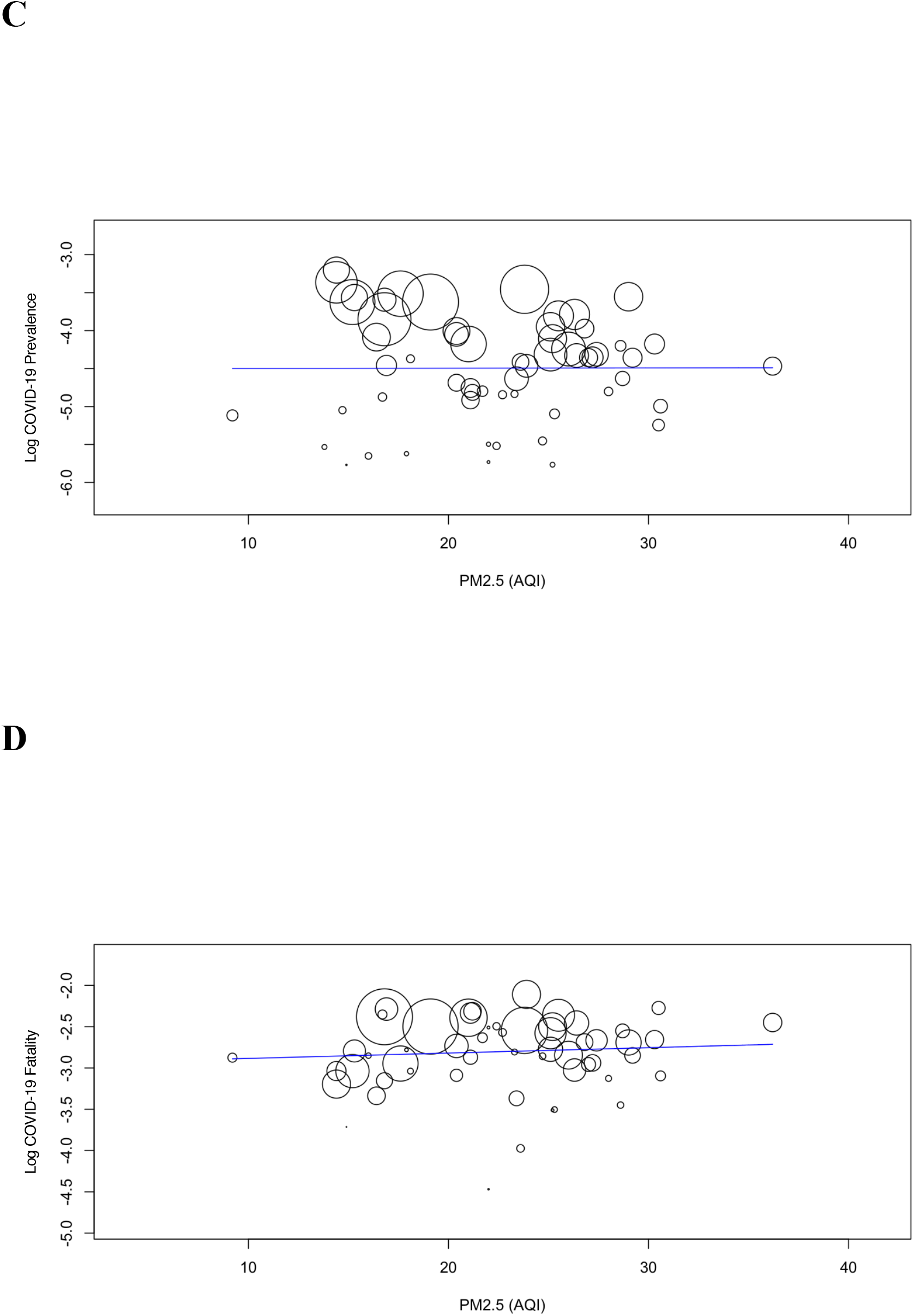
A Meta-Regression Line with Its 95% Confidence-Interval Curves Depicting the COVID-19 Prevalence/Fatality (Plotted as the Logarithm-Transformed Prevalence/Fatality on the Y-Axis) as a Function of Mean Ozone/PM2.5 AQI During 3/15/2020–5/31/2020 (Plotted on the X-Axis).

## Discussion

The present meta-regression suggests no association of real-time/observed ozone/PM2.5 with COVID-19 prevalence/fatality, which is incompatible with previous findings in Chinese 3 cities (Wuhan/XiaoGan/HuangGang),^1^ California state,^2^ Milan metropolitan area (Lombardy, Italy),^3^ and Queens county (NY).^4^ Our analysis included far greater cases (>780,000) and deaths (>52,000) (among >48-million population in the Northeast megalopolis consisting of Boston/New York/Philadelphia/Baltimore/DC/etc.) than these studies.^1-4^ A preliminary study (https://www.medrxiv.org/content/10.1101/2020.04.05.20054502v2) from Harvard University analyzing the largest cases/deaths from >3,000 US counties demonstrated that a small increase in PM2.5 was associated with a large increase in fatality. The investigators, however, used long-term (2000–2016)/estimated PM2.5. In the present study, real-time (3/15/2020–5/31/2020)/observed (neither historical nor estimated) ozone/PM2.5 was analyzed.

In conclusion, ozone/PM2.5 may be unassociated with COVID-19 prevalence/fatality, which should be verified by further experimental/clinical/epidemiological investigations.

## Data Availability

The datasets generated during and/or analysed during the current study are available from the corresponding author on reasonable request.

---

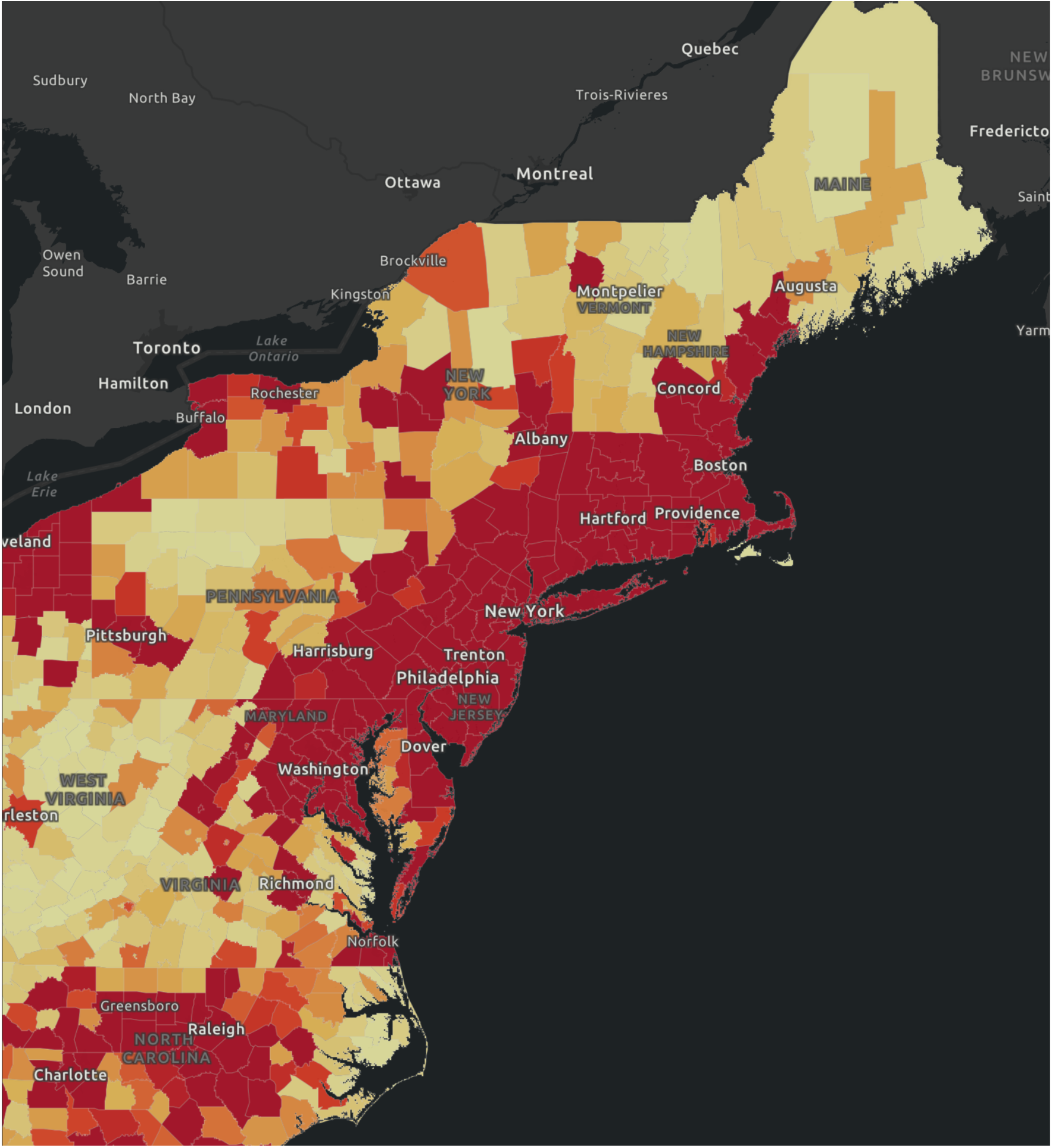
COVID-19 Pandemic in the Northeast Megalopolis (https://coronavirus.jhu.edu/us-map).

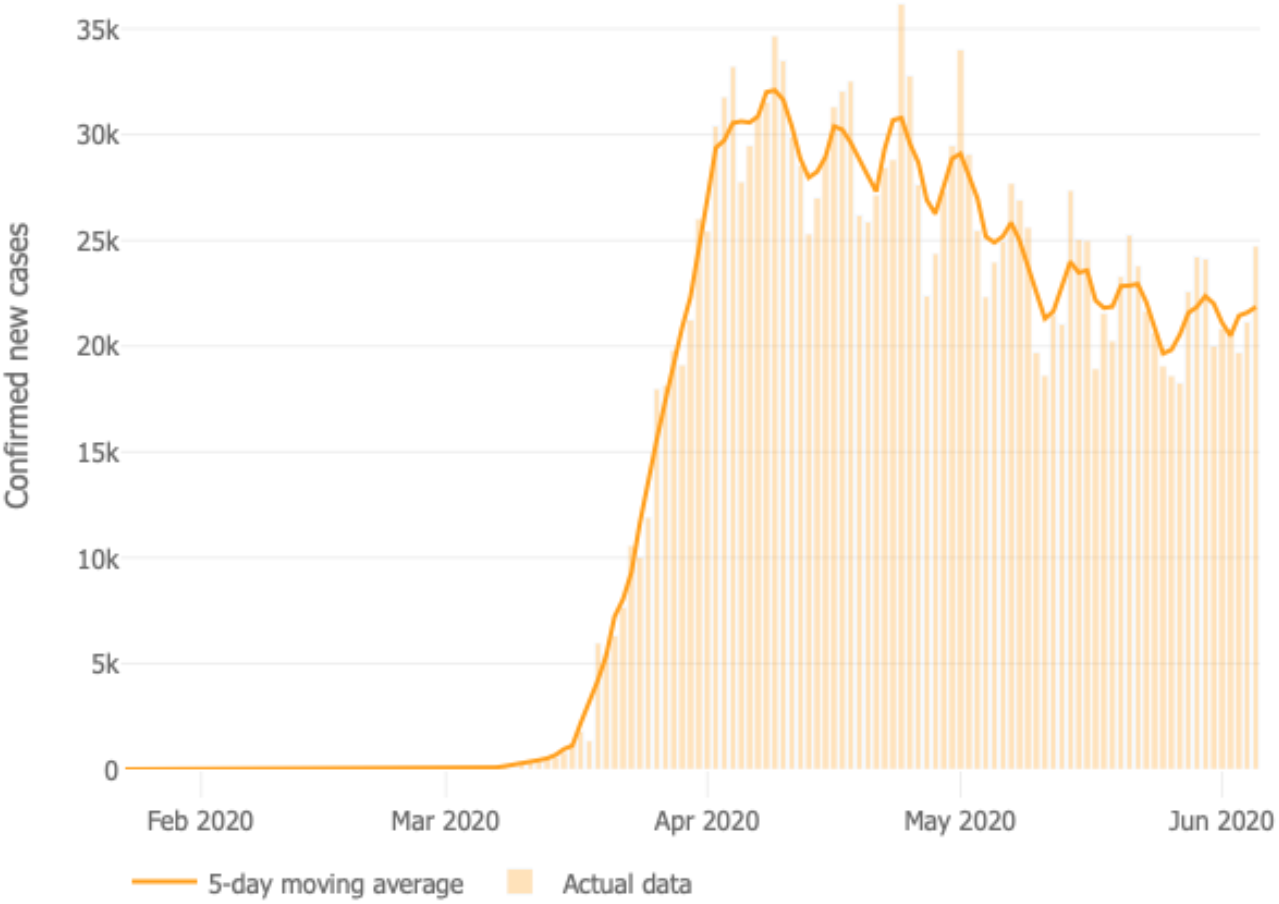
Daily Confirmed New Cases in US (https://coronavirus.jhu.edu/data/new-cases).

